# Validation and reliability of the self-efficacy scale to assess the professional competence of pediatricians participating in continuing medical training program according to the ECHO model

**DOI:** 10.1101/2023.08.03.23293598

**Authors:** Le Hong Nhung, Pham Thu Hang, Nguyen Thu Thuy, Pham Van Hoc, Nguyen Phuong Lan, Pham Duc Han, Piter Martinez Benitez

## Abstract

**Background:** ECHO, (Extension for Community Healthcare Outcomes) model, is a platform for tele-education to expand healthcare workforces to meet the demand of community’s healthcare (1). This program used the connection model between the expert at hubs and the primary health workers (PHWs) in different regions at spokes through video teleconferencing sessions (2). The ECHO program is regarded as a continuous medical training activity to improve professional capacity and job satisfaction at spokes. Currently, there have been many studies on this model, proving that it is effective in improving knowledge and skills for PHWs (3, 4). One of the measures to evaluate the professional development of PHWs is the scale of self-efficacy or self-assessment of competency (5).We carried out a research topic: *"**Validation and reliability of the self-efficacy scale to assess the professional competence of pediatricians participating in continuing medical training according to the ECHO model**"* to help applying to research at ECHO-clinics

**Methods:** Applying cross-sectional study design, implementing mixed methods including quantitative and qualitative in order to adjust the scale of self-efficacy to assess the professional capacity improvement of the healthcare workers to ensure the validity and reliability of the scale. The study proceeds in the following steps: 1) Reviewing the sets of scales for self-assessment of competency (self-efficacy) according to the ECHO model in the world to identify issues of professional competence being assessed and developed the toolkit that suitable to the Vietnamese context; 2) Collecting the decissions from the panel of experts to determine the content of primary toolkit to improve professional capacity through continuous medical training (2); 3) Test the scale on a sample of learners participating in a continuing medical training program using the ECHO model. In this step 3, the research team evaluate the surface validity, the content of validity including the convergent value and discriminant value and the structural reliability with Crobach’s Alpha internal consistency index.

**Results:** The study has reviewed literature about the referred scale of evaluation the professional capacity improvement of health workers following the ECHO model in the world. In order to implement the ECHO model effectively in Vietnam, the study has shown that the adaptation the referred scales need to be dealt in accordance with the socio-cultural-politic context in Vietnam. The adjusted scale of 22 sub-items was built based on the results of qualitative and quantitative research, is considered to be a reliable scale to be applied to the further studies on the ECHO model in Vietnam.

**Conclusions:** The scale to access the healthcare providers’s improvement of professional capacity has been adjusted accordingly to each ECHO program in the world since 2007. However, in this topic, we still conduct the assessment of the level of professional competence. The reliability and validity of the scale according to Delphi and Campell Fiske method to evaluate the face validity and content validity, combined with testing the toolkit in the field to calculate reliability with Crobach Alpha index. Therefore, this assessment tool is a valid and highly reliable.

The scale can be widely applied in evaluating the capacity improvement of PHPs participating in the continuing medical training courses via ECHO model between the hub (the National Children’s Hospital) and the spokes (province-level hospitals in the North of Vietnam). In our research, this scale was built for general uses for any courses with different specialties, the technical terms were adapted to an ECHO program for Continuing Medical Education faculty (CME).

## Background

ECHO, the Extension for Community Healthcare Outcomes (Extension for Community Healthcare Outcomes), is a platform for academic health centers to expand healthcare worker capacity to underserved populations to meet the need of medical healthcare service (2). This program used the model of “hub and spoke” through video conferencing platform to connect the experts at VNCH (hub) with the healthcare workers at different provincial hospital (spoke) (6). The ECHO model has been developed and replicated in many different countries to address health problems and treat common and complex medical conditions, such as HIV/AIDS, substance use disorders, palliative care, rheumatism, chronic diseases and other diseases (7-13). During the implementation process, the ECHO model is regarded as a continuing medical education to improve the professional capacity and professional collaboration ability of health workers in practicum.

Self-efficacy or self-assessment of competency is a concept that has related to the development of medical training since the 90s, in which shows the desire of healthcare workers to make self-determination and self-regulation based on the reality in the treatment process of patients. This desirability depends on the readiness of healthcare workers to self-assess their knowledge, skill and practice based on patient outcomes (5).

Self-assessment of competency in continuing medical education has been fully approved by the American Medical Association (AMA) in its program of retention of practice certificates (14). To evaluate physician’s competence confidence and professional collaboration in clinical practice in the workplace are significant goals in studies to assess the effectiveness of the ECHO model according to Moore’s method (4, 15-17). It is hoped that the results of scale development will help future studies related to the ECHO model with validity and reliability.

The first ECHO model of hepatitis C initiated by Sanjeev used the self-assessment of competency scale named "self-efficacy", laying the foundation for the construction of the scale later in the authors’ studies on different topics. (1) The theoretical basis of the self-efficacy scale included the combination of social cognitive theories of Bandura (18), Vygotsky’s situation-based learning theory (19) and the practice community of Lave (20). The term of “self-efficacy” refers to self-confidence or the perception that one has the ability to organize and take the actions necessary to succeed when given a task.” (Bandura, 1997) (21). Self-efficacy is based on four sources of information: individual’s previous experience, observational experiences in practice, and verbal persuasion on the psychological status of learner. (22)

Later, there had developed many ECHO models, but all were based on these three theories. A number of studies have shown that synergies in learning, coaching and mentoring by experts, among colleagues, have formed “learning loops” (23). In which, emphasis was placed on a concept of self-efficacy assessment at three stages before training, after training and after 3-6 months of training (3) and later also mentioned in other studies and used the concept of “self-assessment of competence” that Masi C developed for the ECHO model of hypertension (24).

When studying ECHO models from the perspective of a continuing medical education (CME) program, the team found that the scale of "self-efficacy" or "self-assessment of competence" had also been developed and applied in different aspects in the implementation process: such as assessing participants’ satisfaction with being able to coordinate and organize the implementation that could run medical service at workplace was known as the scale of “job satisfaction” or “professional satisfaction”

## Methods

### Study setting

The study was conducted at the Vietnam National Children’s Hospital and its satellite hospitals. The study proposal was approved by research ethics committees at the Hanoi University of Public Health (261/2020/YTCCHD3) and the Vietnam National Children’s Hospital (883/BVNTW-VNCSKKTE). The Vietnam National Children’s Hospital was one of the leading pediatric hospitals in Vietnam, located in the North of Vietnam. The hospital played the role as a direction center and provided a training system for satellite hospitals in pediatrics. The direction function of the hospital was represented in the task of updating professional flow and treatment guidelines for satellite hospitals in pediatrics, organizing training courses to update knowledge, and supporting professional resource at distant areas in specific cases. Eventhough, the Vietnam National Children’s Hospital had accomplished certain achievements in providing professional support to the provincial level hospital, the professional support activities were not organized regularly due to limited personel resources, especially in remote areas.

The Project ECHO for pediatricians has been launched since 2019 to improve the capacity of healthcare providers at provincial-level hospitals, then the program officially recruited participants and organized online courses since the beginning of 2020. The anatomy of each session comprised two parts: the theoretical part (didactics) and the practical part (case study discussion). The online course included 8 to 10 sessions depending on the specific course, in which each session lasted about 2 hours with 30 minutes for theoretical presentation and the remaining 90 minutes for discussing a case. A typical online course was conducted in twice per week for 3-5 weeks. In this study, we only focused on learners who participated in the ECHO-Immunology course in 2020. The training hub was Vietnam National Children’s Hospital and the spokes were 18 satellite hospitals which sent learners to participate in the ECHO course. The learners did not pay fees for the courses during this time of study.

## Study design and sampling

Applying cross-sectional research design, using a mix method of qualitative and quantitative research.

### Qualitative study to adjust the scale

***Stage 1:*** Set up the focus group discussion was deployed to find a consensus in the research group (internal concensus) on the origin scale of after translating into Vietnamese, (6) selecting items to form the new scale based on actual implementation including objectives and users.

***Stage 2:*** Invited 06 experts on the CME implementation as panelists to support opinions on the scale:

- The research team called or met in person, wrote emails to discuss with the expert about the purpose of collecting expert’s opinion, and instructed the expert to focus on the content of items in the scale (Appendix 1)
- Collected comments from 06 experts and continued to discuss in the research team, revise the items of scale according to the expert’s comments in each round. The submission was done in 3 rounds for 06 experts with the opinions of all experts in each round.
- Calculated the concensus rate (surface valid) until it reaches above 0.78 which is satisfactory and the collecting expert’s comment process can be stopped. (25)

### Quantitative study to test the adjusted scale in the field

- Choosed the convenient sample with 38 learners who joined in the ECHO course at VNCH.

- Invited them to participate in the study with the answers to the questionaires of the scale, collected feedbacks from learners who were evaluated by the data.

- Calculation of internal consistency reliability Crobach’s Alpha, Convergent value, Discriminant value, some initial results applying the toolkit.

### Study tools and data collection

### Qualitative study tool

Collect expert’s opinion based on Delphi method following the issues:

□ Experts commented on whether the items/sub-items are appropriate to measure the “self-assessment of competency” before and after participating the online education program following ECHO model.

□ Experts make any possible suggestions for adding or removing items or changing the wording of items on the scale.

□ Experts evaluate the instruction of scale.

□ Experts evaluate the format of the scale.

### Quantitative study tool

The questionnaire was built after 3 rounds of expert opinion and was adjusted based on the feedbacks between periods, the experts were asked to give a mark from 1 to 4 on a Likert scale with level 1: Very unsuitable; level 2: Not suitable; level 3: Suitable; level 4: Very suitable.

The initial questionaire after being translated from the original questionnaire from Sanjeev Arora consisted of 34 questions, then adjusted based on internal discussions in the research group that include 28 questions: self-assessment of the physician’s professional competence, divided into 4 major categories including part 1: self-assessment of competence in knowledge and professional practice before training, including 6 questions; scale 2: self-assessment of competence in knowledge and professional practice after training includes 6 questions; scale 3: self-assessment of professional satisfaction in clinical practice before training includes 8 questions; scale 4: self-assessment of professional satisfaction in clinical practice after training includes 8 questions.

### Data collection and analysis

The guideline for collecting expert opinions was carried out 3 times based on the concensus rate and the median value in each period to select sub-items.

- Quantitative scale was built and sent to the participating experts for rating from 1 to 4 point to show the degree of agreement. With the point 3 and 4 obtained, the research team would calculate the concensus rate (surface value) after consultations until it reached above 0.78, then stoped collecting the expert’s opinions.
- The final adjusted scale with high approval would be tested a reliability using Crobach’s Alpha internal consistency index and validated with the content including the Convergent and Discriminant value following Campell&Fiske method.

### Result and Discussion

### Stage 1 and 2: Developing invitation form and sending the invitation to experts for collecting opinion

Based on the literature review, we obtained an initial set of 34 questions in the scale of self-assessment of competency (Table 1 and Table 2).

**Table 1:** Specialists’ characteristics.

| **Characteristics** | **N(%)** | **(Mean, SD)** |
| --- | --- | --- |
| ***Total*** | **6** |  |
| ***Sex*** |  |  |
| Male | 3 (50%) |  |
| 0%) Female | 3 (50%) |  |
| ***Title*** |  |  |
| Master | 2 (33,3%) |  |
| PhD | 3 (50%) |  |
| Asso.Professor | 1 (16,7%) |  |
| ***Years of working experience*** |  | 16,2 (4,2) |

**Table 2:** the initial scale.

|  |
| --- |
| ***Part I: Scale evaluate the self-assessment of competency in knowledge, professional practice following Likert 5 before training***<br>(Learner tick the point from 1 to 5 under each question 1:Very poor;2:Poor; 3: Average;4: Good; 5: Excellent) |
| **Sentence 1:** *Ability to detect symptoms of patients who need to be screened* |
| **Sentence 2:** *Ability to detect patients suitable for treatment?* |
| **Sentence 3:** *Ability to assess the extent of damage to the related organs in the patient* |
| **Sentence 4:** *Ability to treat patients and manage side effects* |
| **Sentence 5:** *Ability to educate and motivate patients* |
| **Sentence 6:** *Ability to serve as a consultant in the clinic and in local area* |
| ***Part II: Scale evaluate the self-assessment of competency in knowledge, professional practice following Likert 5 after training***<br>(Learner tick point from 1 to 5 under each question 1:Very poor ;2:Poor; 3: Average;4: Good; 5: Excellent) |
| **Sentence 1:** *Ability to detect symptoms of patients who need to be screened* |
| **Sentence 2:** *Ability to detect patients suitable for treatment* |
| **Sentence 3:** *Ability to assess the extent of damage to the related organs in the patient* |
| **Sentence 4:** *Ability to treat patients and manage side effects* |
| **Sentence 5:** *Ability to educate and motivate patients* |
| **Sentence 6:** *Ability to serve as a consultant in the clinic and in local area* |
| ***Part III: Scale evaluate the the self-efficacy of job satisfaction following Likert 5 before training***<br>(Learner tick point from 1 to 5 under each question 1:Very poor ;2:Poor; 3: Average;4: Good; 5: Excellent) |
| **Sentence 1:** *I feel professionally isolated at work* |
| **Sentence 2:** *I can create relationships easily with colleagues* |
| **Sentence 3:** *I easily reach out to my doctor if I need professional feedback or help from them* |
| **Sentence 4:** *I easily access resources for career development* |
| **Sentence 5:** *When I need help and support from a doctor, I can contact an expert at the appropriate time* |
| **Sentence 6:** *I have the opportunity to regularly share my clinical experience with my colleagues* |
| **Sentence 7:** *In general, I am satisfied with the job* |
| **Sentence 8:** *I am sure that I can improve the quality of medical examination and treatment services in my specialty.* |
| **Part IV: Scale evaluate the the self-efficacy of job satisfaction following Likert 5 after training**<br>(Learner tick point from 1 to 5 under each question 1:Very poor ;2:Poor; 3: Average;4: Good; 5: Excellent) |
| **Sentence 1:** *I feel professionally isolated at work* |
| **Sentence 2:** *I can create relationships easily with colleagues* |
| **Sentence 3:** *I easily reach out to my doctor if I need professional feedback or help from them* |
| **Sentence 4:** *I easily access resources for career development* |
| **Sentence 5:** *When I need help and support from a doctor, I can contact an expert at the appropriate time* |
| **Sentence 6:** *I have the opportunity to regularly share my clinical experience with my colleagues* |
| **Sentence 7:** *In general, I am satisfied with the job* |
| **Sentence 8:** *I am sure that I can improve the quality of medical examination and treatment services in my specialty.* |

After reviewing and discussing in the research group, we removed items not involved the online course conducted at Vietnam National Children’s Hospital.

Send 06 experts for comments (Details of invitation letter are Appendix 2A). Toolkit sent to experts for the first time includes 28 questions (Detailed Table 2)

#### Stage 3: The results of assessment of 6 experts through roundtable discussions

Taking into consideration the 6 experts’ comments in the first roundtable discussion, the result was as followed:

**Table 3:** the first roundtable discussion result.

| **Section I, II: Scale evaluate the self-assessment of competency in knowledge, professional practice following Likert 5 BEFORE and AFTER training**<br>(Learner tick the point from 1 to 5 under each question 1:Very poor;2:Poor; 3: Average;4: Good; 5: Excellent) | **Median**<br>**(Q1, Q3)** | **Consensus**<br>**rate**<br>**(%)** |
| --- | --- | --- |
| **Sentence 1:** *Have the ability to detect symptoms of patients who need to be screened* | 3,0<br>(2,75,0;3,0) | 83,3% |
| **Sentence 2:** *Have the ability to detect patients suitable for treatment* | 3,0(2,0;3,0) | 66,7% |
| **Sentence 3:** *Have the ability to assess the extent of damage done to relatable organs in patients* | 2,5(1,75;3,00) | 50% |
| **Sentence 4:** *Have the ability to treat the patients and manage one's side effects* | 3,0(2,0;3,0) | 66,7% |
| **Sentence 5:** *Have the ability to educate and motivate the patients* | 2,5(2,0;3,0) | 50% |
| **Sentence 6:** *Have the ability as a consultant in my clinic as well as locally regarding medical problems* | 2,5(2,0;3,0) | 50% |
| **Section III, IV: Scale evaluate the self-efficacy of job satisfaction following Likert 5 BEFORE and AFTER training**<br>(Learner tick the point from 1 to 5 under each question 1: Very poor; 2: Poor; 3: Average; 4: Good; 5: Excellent) |  |  |
| **Sentence 1:** *I feel isolated when working professionally at my workplace* | 2,5(2,0;3,0) | 50% |
| **Sentence 2:** *I could easily form close relationships with my co-workers* | 2,5(1,75-3,0) | 50% |
| **Sentence 3:** *I could easily access a doctor when I needed their professional feedback or their help* | 3,0(2,0; 3,0) | 66,7% |
| **Sentence 4:** *I could easily access all sources and documents for my career development (improvement in my knowledge)* | 2,0(1,0;2,25) | 16,7% |
| **Sentence 5:** *When I needed the clinical doctors' help or support, I was able to contact the experts in due time.* | 3,0 (2,0;3,0) | 66,7% |
| **Sentence 6:** *I was able to have the opportunity to share my clinical experiences with my co-workers at regular intervals* | 3,0(2,00;3,00) | 66,7% |
| **Sentence 7:** *I'm generally satisfied with my job* | 3,0 (2,0;3,0) | 66,7% |
| **Sentence 8:** *I'm confident that I could improve the quality of medical service delivery in my facility* | 3,0(2,75;3,0) | 83,3% |

The consensus rate was calculated as followed: percentage of responses with a score of 3 or 4 applied for all categories regarding the 6 specialists. Based on the quantitative comments, the sentence with a consensus rate lower than 50% would be excluded, and sentence with a consensus rate from 50% - 78% would be continued to rate under consideration by the experts’ detailed comments. In sections I and II, with sentence 1 and sentence 2, sentence 3, it was necessary to consider to clarify the differences between those sentences. Furthermore, experts gave two additional sentences based on the necessity of the doctor’s assignment revolving emergency management: “Ability to handle emergency situations according to standard protocols”. Based on the necessity in prognosis issues and management of associated complications of the main disease, the experts gave an additional sentence “Ability to manage complications of diseases". The content of sentence 6 of sections I and II coincided in 3 sentences, including sentence 5 of sections I and II and sentences 7 and 8 in sections III and IV. Thus, it was necessary to discard two out of three and adjust the technical terms precisely. In sentence 1 of sections III and IV, experts believed that we should change the statement “I feel isolated when working professionally at my workplace” into the statement “Ability to resolves indepently professional issues at the workplace." Besides, sentences 2 and 6 in sections III and IV had an inclusive content, an expert delivered the opinion that should group the sentences into a general sentence "Ability to exchange clinical experiences among colleagues." Sentences 3 and 5 in ***sections III and IV*** need to be grouped as the contents were similar.

In addition to removing and adding words in sentences, all sentences faced the experts’ revising opinion for a more suitable terminology. In sentence 5 of sections I and II, the specialists suggested Vietnameseized uses with the medical terminology: "Ability to advise and educate patients on self-care and disease prevention”.

Therefore, in the second round of discussion, the research team decided to include two additional sentences and asked for all the experts’ comments, removing sentences with a score of less than 50% and need the further revision. The research team synthesized the valuation sheet, presenting the questionnaire of 22 questions was regarded as the scale of self-assessment of competency.

Finally, the scale of self-assessment of competency measured the change of a doctor’s knowledge and professional practice before and after being trained, which consisted of four sections. Section I*: " Scale evaluate the self-assessment of competency in knowledge, professional practice following Likert 5 BEFORE training"* consists of 6 questions. Section II*: "Scale evaluate the self-assessment of competency in knowledge, professional practice following Likert 5 AFTER training* consists of 6 questions. Section III: *" **Scale evaluate the self-efficacy of job satisfaction following Likert 5 BEFORE training** "* consisted of 5 questions, and section IV*: " **Scale evaluate the self-efficacy of job satisfaction following Likert 5 AFTER training** "*consists of 5 questions.

Regarding the layout of the scale, all experts agreed upon using the Likert 5 scale. In order to stay in line with the course framework’s goals and adjust the medical terminology, the revised scale was continued to send to the six experts in round 2 for revision.

**Table 4.** the second roundtable discussion result.

| *Section I, II: Scale evaluate the self-assessment of competency in knowledge, professional practice following Likert 5 BEFORE and AFTER training*<br>*(Learner tick the point from 1 to 5 under each question*<br>*1:Very poor;2:Poor; 3: Average;4: Good; 5: Excellent)* | *Median*<br>*(Q1, Q3)* | *Consensus rate*<br>*(%)* |
| --- | --- | --- |
| Sentence 1: Ability to detect symptoms of patients who need to be screened. | 3,00 (3,00;3,00) | 100% |
| Sentence 2: Ability to analyze and synthesize clinical and subclinical manifestation to make an appropriate diagnosis. | 3,00(2,75;4,00) | 83,3% |
| Sentence 3: Ability to apply standardized treatment protocol and control side effects when prescribing. | 3,50(2,75;4,00) | 83,3% |
| Sentence 4: Ability to handle emergency situations according to standard protocols. | 3,00(2,75; 3,25) | 83,3% |
| Sentence 5: Ability to advise and educate patients on self-care and disease prevention. | 3,50 (2,75;4,00) | 83,3% |
| Sentence 6: Ability to manage complications of the disease. | 3,00(2,00;3,00) | 66,7% |

| **Section III, IV: Scale evaluate the self-efficacy of job satisfaction following Likert 5 BEFORE and AFTER training**<br>*(Learner tick the point from 1 to 5 under each question 1: Very poor; 2: Poor; 3: Average; 4: Good; 5: Excellent)* | **Median**<br>**(Q1, Q3)** | **Approval rate**<br>**(%)** |
| --- | --- | --- |
| Sentence 1: Ability to resolves indepently professional issues at the workplace. | 3,00(2,75;3,25) | 83,3% |
| Sentence 2: Ability to exchange clinical experiences among colleagues. | 3,00(2,75;3,00) | 83,3% |
| Sentence 3: Ability to get experts' support to solve illness cases at the workplace. | 3,00(2,75;3,25) | 83,3% |
| Sentence 4: Ability to finish the assigned work according to one's expertise in the workplace. | 3,00(2,75;4,00) | 83,3% |
| Sentence 5: Ability to improve the quality of the treatment and examination services according to one's professional department in the workplace. | 3,0(2,75;4,00) | 83,3% |

Based on the quantitative comments, sentences that had a consensus rate at least 78% would be chosen from the expert’s assessment panel. The result of the second round included 22 questions with 6 couple of questions regarding confidence in professional knowledge and practical expertise before and after training, and 5 couple of questions regarding one’s ability to coordinate professionally in clinical practices. An expert commented to change sentence 6 in sections I and II into the sentence of *"Ability to manage the risks involved, detect and smartly solve the complications of the illness."*

Regarding the measurement format, one expert add the instruction placed after the name of the scales that “students can skip this section if they deem it unsuitable”. In addition, it was necessary to divide the columns as BEFORE training and AFTER training because of the same contents in scales I and II, and scales III and IV. Thus, the final scale in round 2 had 2 sections with the Section I: “*Scale evaluate the self-assessment of competency in knowledge, professional practice following Likert 5 BEFORE and AFTER training”* and the section II: “ *Scale evaluate the self-efficacy of job satisfaction following Likert 5 BEFORE and AFTER training”,* as followed the table 5.

**Table 5:**
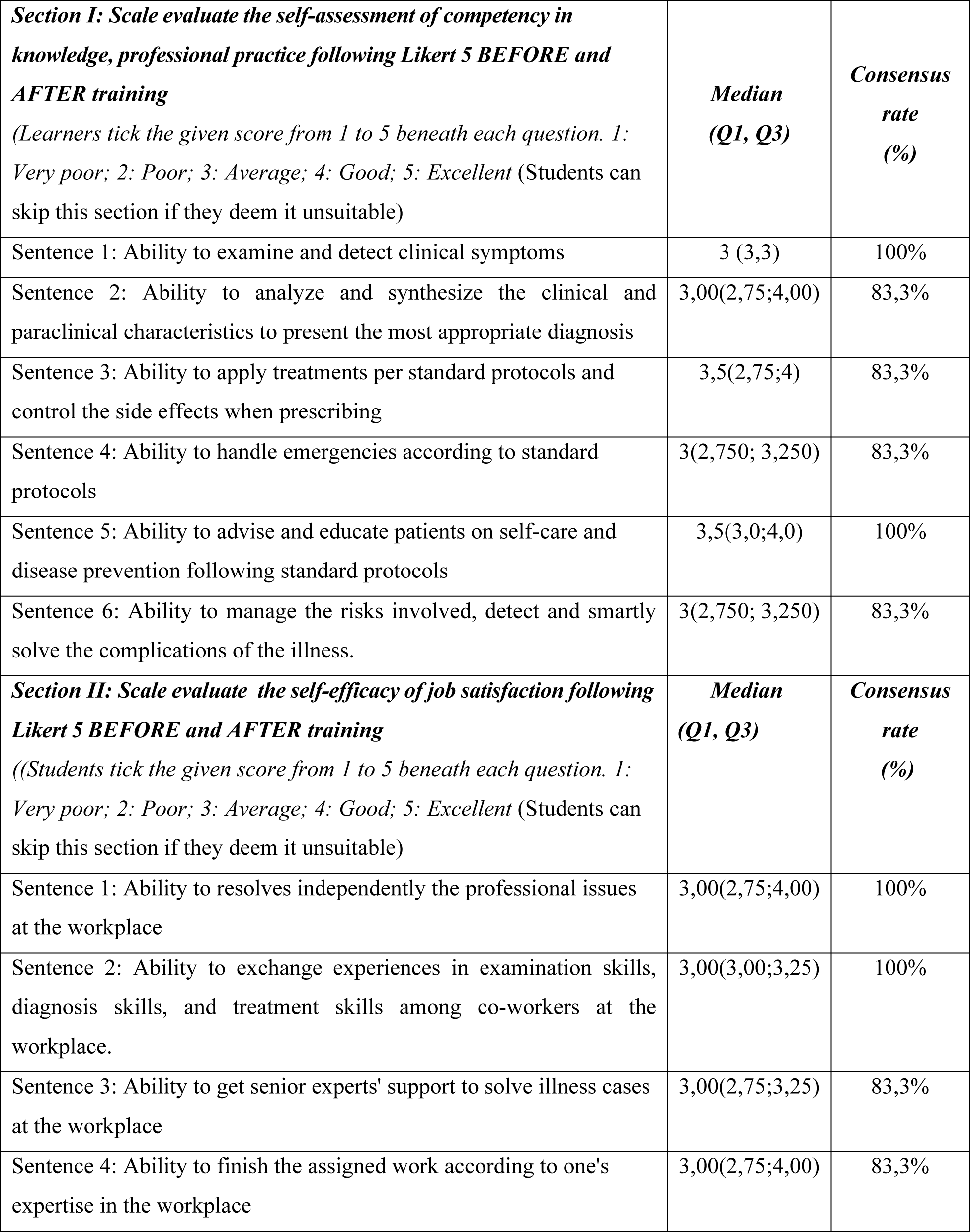
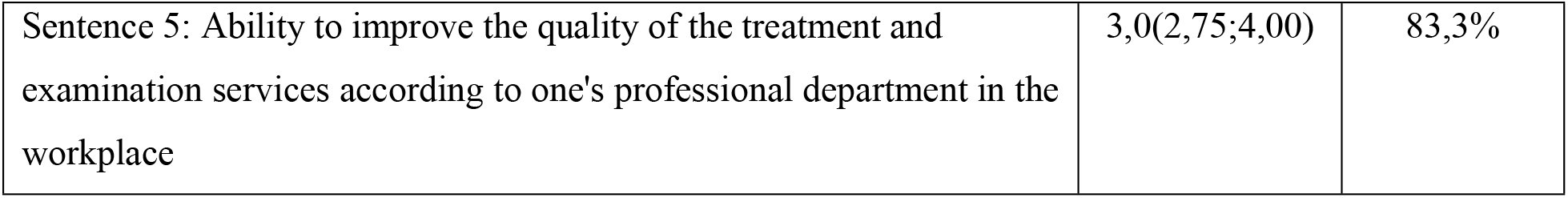
the third roundtable discussion.

|  |  |  |
| --- | --- | --- |
| **Section I: Scale evaluate the self-assessment of competency in knowledge, professional practice following Likert 5 BEFORE and AFTER training**<br>*(Learners tick the given score from 1 to 5 beneath each question. 1: Very poor; 2: Poor; 3: Average; 4: Good; 5: Excellent (Students can skip this section if they deem it unsuitable)* | **Median**<br>**(Q1, Q3)** | **Consensus**<br>**rate**<br>**(%)** |
| Sentence 1: Ability to examine and detect clinical symptoms | 3 (3,3) | 100% |
| Sentence 2: Ability to analyze and synthesize the clinical and paraclinical characteristics to present the most appropriate diagnosis | 3,00(2,75;4,00) | 83,3% |
| Sentence 3: Ability to apply treatments per standard protocols and control the side effects when prescribing | 3,5(2,75;4) | 83,3% |
| Sentence 4: Ability to handle emergencies according to standard protocols | 3(2,750; 3,250) | 83,3% |
| Sentence 5: Ability to advise and educate patients on self-care and disease prevention following standard protocols | 3,5(3,0;4,0) | 100% |
| Sentence 6: Ability to manage the risks involved, detect and smartly solve the complications of the illness. | 3(2,750; 3,250) | 83,3% |
| **Section II: Scale evaluate the self-efficacy of job satisfaction following Likert 5 BEFORE and AFTER training**<br>*((Students tick the given score from 1 to 5 beneath each question. 1: Very poor; 2: Poor; 3: Average; 4: Good; 5: Excellent (Students can skip this section if they deem it unsuitable)* | **Median**<br>**(Q1, Q3)** | **Consensus**<br>**rate**<br>**(%)** |
| Sentence 1: Ability to resolves independently the professional issues at the workplace | 3,00(2,75;4,00) | 100% |
| Sentence 2: Ability to exchange experiences in examination skills, diagnosis skills, and treatment skills among co-workers at the workplace. | 3,00(3,00;3,25) | 100% |
| Sentence 3: Ability to get senior experts' support to solve illness cases at the workplace | 3,00(2,75;3,25) | 83,3% |
| Sentence 4: Ability to finish the assigned work according to one's expertise in the workplace | 3,00(2,75;4,00) | 83,3% |
| Sentence 5: Ability to improve the quality of the treatment and examination services according to one's professional department in the workplace | 3,0(2,75;4,00) | 83,3% |

According to Lawshe’s suggestion, consensus rate that higher than 0.78 were considered satisfactory (25). The research team had already carefully examined the final scale based on the three times of the feedbacks from experts. Sections were combined, the unclear questions were removed.

**Table 6:** The final scale.

|  |  |  |
| --- | --- | --- |
| ***Section I: Scale evaluate the self-assessment of competency in knowledge, professional practice following Likert 5.***<br>*((Students tick the given score from 1 to 5 beneath each question. 1: Very poor; 2: Poor; 3: Average; 4: Good; 5: Excellent (Students can skip this section if they deem it unsuitable))* | ***BEFORE***<br>***training*** | ***AFTER***<br>***training*** |
| Sentence 1: Ability to examine and detect clinical symptoms. |  |  |
| Sentence 2: Ability to analyze and synthesize the clinical and paraclinical characteristics to present the most appropriate diagnosis. |  |  |
| Sentence 3: Ability to apply treatments per standard protocols and control the side effects when prescribing. |  |  |
| Sentence 4: Ability to handle emergencies according to standard protocol(s). |  |  |
| Sentence 5: Ability to manage the risks involved, detect and solve the complications of the illness. |  |  |
| Sentence 6: Ability to advise and educate patients on self-care and disease prevention following standard protocol(s). |  |  |
| ***Section II: Scale evaluate the self-efficacy of job satisfaction following Likert 5***<br>*((Students tick the given score from 1 to 5 beneath each question. 1: Very poor; 2: Poor; 3: Average; 4: Good; 5: Excellent (Students can skip this section if they deem it unsuitable))* | ***BEFORE***<br>***training*** | ***AFTER***<br>***training*** |
| Sentence 1: Ability to independently resolves professional issues at the workplace. |  |  |
| Sentence 2: Ability to exchange experiences in examination skills, diagnosis skills, and treatment skills among co-workers at the workplace. |  |  |
| Sentence 3: Ability to get senior experts' support to solve illness cases at the workplace. |  |  |
| Sentence 4: Ability to finish the assigned work according to one's expertise in the workplace. |  |  |
| Sentence 5: Ability to improve the quality of the treatment and examination services according to one's professional department in the workplace. |  |  |

#### Stage 4: Field test result

The final scale (22 items) was distributed by the research team to all learners participating in the online training program following the ECHO-Immunology course at the National Children’s Hospital. None of the learners were disqualified. The survey was distributed to learners in an online session after the end of the course. tooltkit guidelined the instruction on the objective of the questionnaire, the way of answering questions, and ensuring anonymity. The questions also included demographic characteristics, the name of the program, and the learner’s qualifications when participating in the program.

**Table 6:** Characteristics of learners participating in the training program.

| *Characteristics* | *N (%)* |
| --- | --- |
| ***Total*** | **38** |
| ***Gender*** |  |
| Male | 10 (26,3%) |
| Female | 28 (73,7%) |
| ***Profession*** |  |
| General Pediatrician | 22 (58%) |
| Master of Physician | 11 (29%) |
| Fellow of Pediatrician/PhD | 5 (13%) |
| ***Year numbers of working*** |  |
| < 5 | 8 (21,1%) |
| 5-9 | 10 (26,3%) |
| 10-14 | 11 (28,9%) |
| >= 15 | 9 (23,7%) |

### The results of validity and reliability of the scale

The quality and completeness of the feedbacks from 38 learners response to the scale were assessed. The mean and standard deviation were calculated based on the points in each item category of 38 learners. The calculated percentage for missing items or domains with accepted values were less than 20%. A high number of missing items or a high percentage of missing data throughout the questionnaire indicated that the items were confusing or that the questionnaire/scale layout was problematic (26). Spearman’s correlation was used in this study with a non-normal distribution.

**Table 7:** Characteristics of item statistics.

|  | **Items** | **Missing value (%)** | **Mean interval** | **Standard deviation Interval (SD)** |
| --- | --- | --- | --- | --- |
| *Self-assessment of competency in knowledge, professional practice before training* | 6 | 0 | 2.18-2.37 | 0.68-0.75 |
| *Self-assessment of competency in knowledge, professional practice after training* | 6 | 0 | 3.55-3.68 | 0.47-0.6 |
| *Self-efficacy of job satisfaction before training* | 5 | 0 | 2.37-2.55 | 0.82-0.47 |
| *Self-efficacy of job satisfaction after training* | 5 | 0 | 3.45-3.63 | 0.49-0.56 |

The mean score and standard deviations of the items in each domain before training ranged from 2.18±0.68 to 2.55±0.47; The mean score and standard deviation of the items in each domain after training ranged from 3.45±0.49 to 3.68±0.6.

### The construct validity including convergent and discriminant validity

The construct validity is evaluated by calculating the item’s convergent and discriminant validity. The correlation of each item with its own total score was considered to be satisfactory if it valued > 0.30 (27). Theoretically, the convergence value in the same item was higher than the correlation value in other items. The discriminant validity of the item assumed that in the tool with more than one domain, the correlations between items in the same domain were expected to be higher significantly than those in other domains. The scale success rate was calculated, as suggested by McHorneys et al., (28) as the percentage of items in each domain that met the criteria for convergent validity and discriminant validity. Following the matrix model Campell & Fiske, the result was interpreted at the diagonal values.

**Table 8:**
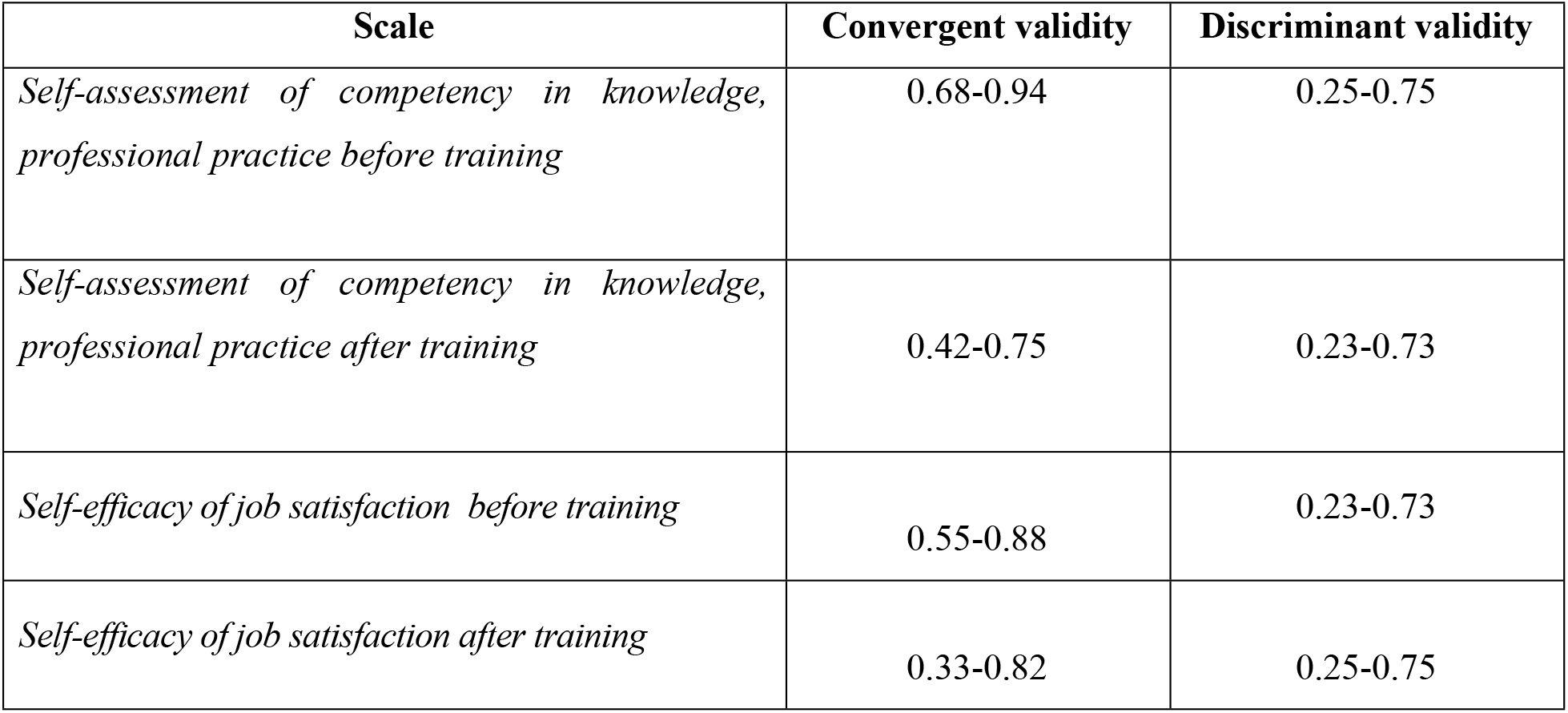
results of multi-point matrix correlation.

| **Scale** | **Convergent validity** | **Discriminant validity** |
| --- | --- | --- |
| *Self-assessment of competency in knowledge, professional practice before training* | 0.68-0.94 | 0.25-0.75 |
| *Self-assessment of competency in knowledge, professional practice after training* | 0.42-0.75 | 0.23-0.73 |
| *Self-efficacy of job satisfaction before training* | 0.55-0.88 | 0.23-0.73 |
| *Self-efficacy of job satisfaction after training* | 0.33-0.82 | 0.25-0.75 |

#### Internal consistency reliability

Internal consistency reliability was tested by Crobach’s Alpha coefficient for each domain and for the whole toolkit with an acceptable value of Crobah Alpha ≥ 0.70. However, if Crobach Alpha value ≥ 0.60 was considered acceptable in the newly developed scale (29). Cronbach’s Alpha was also checked when individual items were deleted. Items that reduced the Cronbach’s Alpha value in the domain were considered for exclusion.

**Table 9:** Crobach Alpha reliability.

|  | **Inter-scale correlation** |  |  |  | **Crobach Alpha** |
| --- | --- | --- | --- | --- | --- |
|  | *Self-assessment of competency in knowledge, professional practice before training* | *Self-assessment of competency in knowledge, professional practice after training* | *Self-efficacy in professional collaboration in professional practice before training* | *Self-efficacy in professional collaboration in professional practice after training* |  |
| *Self-assessment of competency in knowledge, professional practice before training* | 1 | - | - | - | 0,829 |
| *Self-assessment of competency in knowledge, professional practice after training* | 0,744 | 1 | 0,72 | - | 0,818 |
| *Self-efficacy of job satisfaction before training* | 0,789 | - | 1 | - | 0,84 |
| *Self-efficacy of job satisfaction after training* | 0,764 | 0,84 | 0,813 | 1 | 0,875 |

To further test whether the domains were measured the different aspects of professional competence confidence, the Cronbach’s alpha of each domain section was compared with another domain section correlation coefficient. If Cronbach’s alpha of each domain was higher than the domain correlation indicated that the domain scores represented the different aspects of professional competence confidence. [5]

## Conclusion

The scale to assess the learners’s professional capacity improvement had been applied and adjusted accordingly to each ECHO program in the world since 2007. However, in this topic, we still conducted the scale for the assessment of the level of professional competence. Delphi method was ultilised to evaluate the surface validity, combined with testing the scale in the field in order to calculate the construct or content validity and reliability following Campell & Fiske method. Therefore, this toolkit was a valid and highly reliable scale that would be widely applied in evaluating the impact of continuing medical training courses following ECHO model at Vietnam National Children’s Hospital.

Moreover, thanks to the progress of ECHO model implementation, we recommended that the scale should be built and adjusted based on the disease model of each locality and customised to each organization to implement. In our study, this scale was built for general use in the ECHO courses with different specialties, the medical terminology in the domains were adjusted to fit an ECHO program for medical training continuous purpose.

### List of Abbreviations

AMA: American Medical Association
CME: Continuing Medical Education
CV: Convergent validity
DV: Discriminat validity
ISC: Inter scale correlation
ECHO: Extension for Community Healthcare Outcomes
HCV: Hepatitis C Virus
HIV: Human Immunodeficiency Virus
MOH: Ministry of Health
SD: Standard deviation
Q1, Q2, Q3: quartile
WHO: World Health Organization

## Declarations

### Ethics approval and consent to participate

The study proposal was approved by research ethics committees at the Hanoi University of Public Health (261/2020/YTCC-HD3) and the Vietnam National Children’s Hospital (883/BVNTW-VNCSKKTE). The study was conducted in accordance with relevant guidelines and regulations. Informed consent was obtained from all participants.

### Consent for publication

Not applicable

### Availability of data and materials

The datasets used and/or analysed during the current study are available from the corresponding author on reasonable request.

### Authors’ contributions

LHN, PMB and NPL, PVH developed the study concept and design, wrote the main manuscript text. LHN and PMB contributed to the data collection and analysis. NTT, PTH and PDH contributed to revise and improve the manuscript.

All authors read and approved the final manuscript.

## Data Availability

the full data is available after acceptance. Relevant data are within the manuscript.

## Acknowledgments

The authors thank hospital managers and healthcare providers who participated in this study. The views expressed in this article are solely those of the authors and do not represent the official positions of the organizations the authors are affiliated with.

## APPENDIX 1: INVITATION LETTER FOR EXPERT’S RATING AT THE FIRST ROUNDTABLE DISCUSSION

Within the framework of the study to improve the professional competence and job satisfaction in clinical practice of learners after participating in the Continuing Medical Education program following ECHO model, we invite you to evaluate the scale that we have developed and adjusted. The name of the scale is " self-assessment of learners’ professional competence when participating in the online Continuing Medical Education program". We would like to invite you to evaluate the content and the layout of the scale.

### 1. Evaluation process

□ Experts read the description of the summary of the theoretical basis of scale development
□ Experts use the evaluation form and evaluates each item following the point level from 1 to 4 (1- very unsuitable;2- unsuitable, 3- suitable and 4-very suitable)
□ Experts comment whether the items/sub-items in the scale are appropriate to measure “self-assessment of competency” in knowledge, professional practice and “self-efficacy of job satisfaction” according to the Likert 5 scale before and after participating in the Continuing Medical Education program
□ Experts make any possible suggestions for adding or removing items or changing the wording of items on the scale.
□ Experts evaluate the instruction for the scale.
□ Experts evaluate the layout of the scale.

### 2. Scale Instruction

□ Describe the conceptual framework for scale development.
□ Describe the scale.
□ Form for assessing the appropriatebility of the scale.

#### 2.1. Conceptual framework to develop the scale

The first course followed ECHO model initiated by Arora et al. on hepatitis C used the "self-efficacy" scale, laying the background for the construction of the scale in studies on different professional issues. The theoretical basis of the "self-efficacy" scale consists of a combination of Bandura’s theories of social cognition, Vygotsky’s theory of case-based learning and the community of practice. In particular, emphasizing a concept of "self-efficacy" performance assessment that Prof. Arora had researched to build into the scale to evaluate participants’s learning through online programs at the periods of pre-training, post-training and after 6 months of training and later Jane Wright also developed a set of questionaire to assess coordination ability that would satisfy for the work of primary care physicians participating in the ECHO-HIV program.

##### Description about the adjusted scale

For these scales, we are interested in how physicians’ competence improve and ability to demonstrate expertise at workplace that be evaluated by themselves before and after participation in the course of continuing medical training. After being adjusted and tested at field, the scale will be used in online course feedback (ECHO model course) from learners to find out how they feel and evaluate about Continuing Medical Education. There are a total of 4 scales.

Divided into 2 periods to evaluate including before and after the learners being trained. On the scale of assessment of competency and ability to respond to professional expertise before and after training, it is assessed on a 5 level Likert scale from 1:Very poor;2: Poor; 3: Average; 4: Good; 5: Excellent.

##### The form to assess the approriateness of the initial toolkit

###### Guidance

Please use the following form to assess the appropriateness of each item to the concepts of "self-assessment of competency" and "self-efficacy of job satisfaction”

Please read each item or sentence carefully; then rate each item or sentence on a scale from one to four score, depending on the relevance of result expectation that experts expect.

***“ 1 = very unsuitable”***

***“2= unsuitable”***

***“3= suitable”***

***“4 = very suitable”***

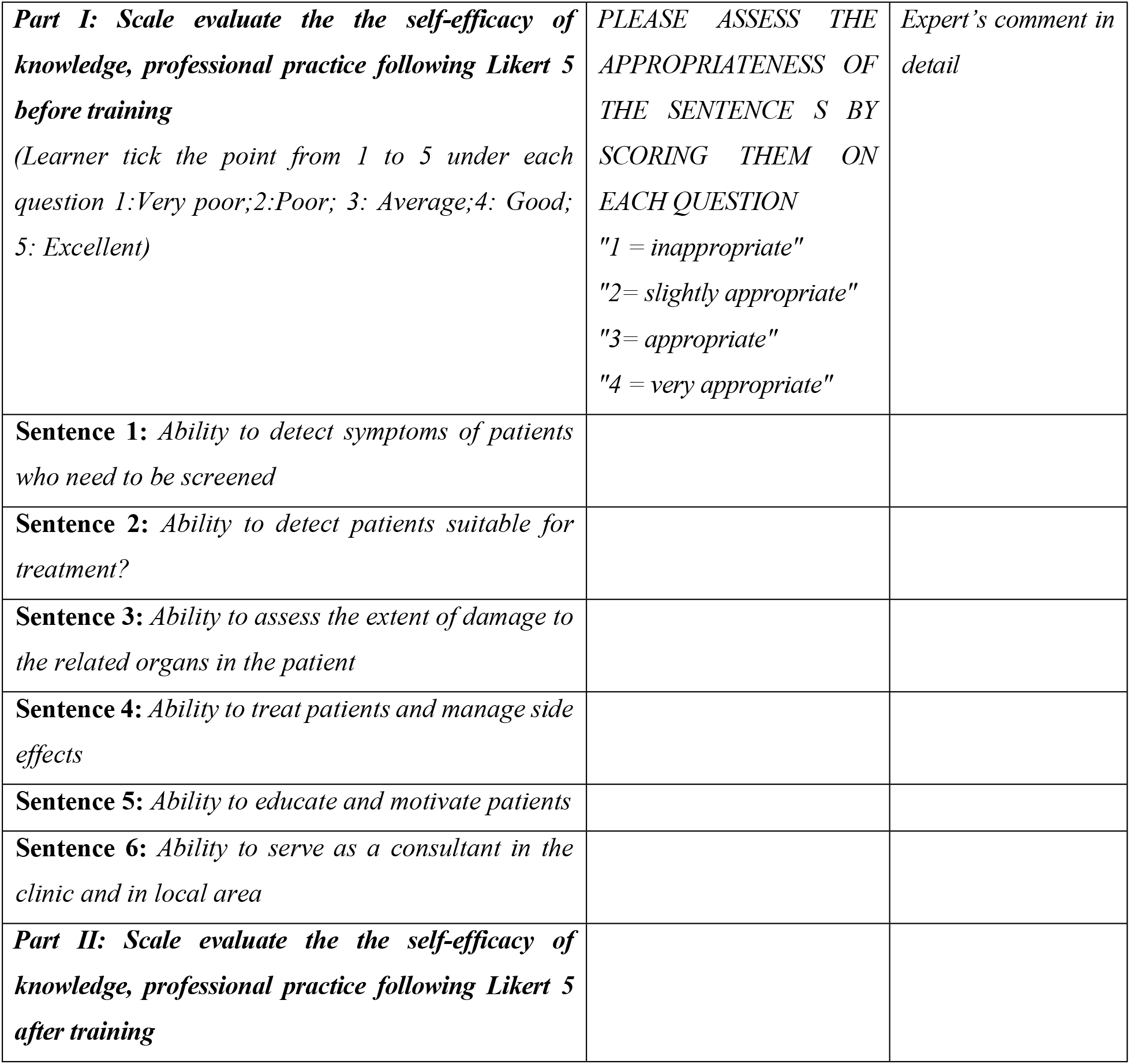
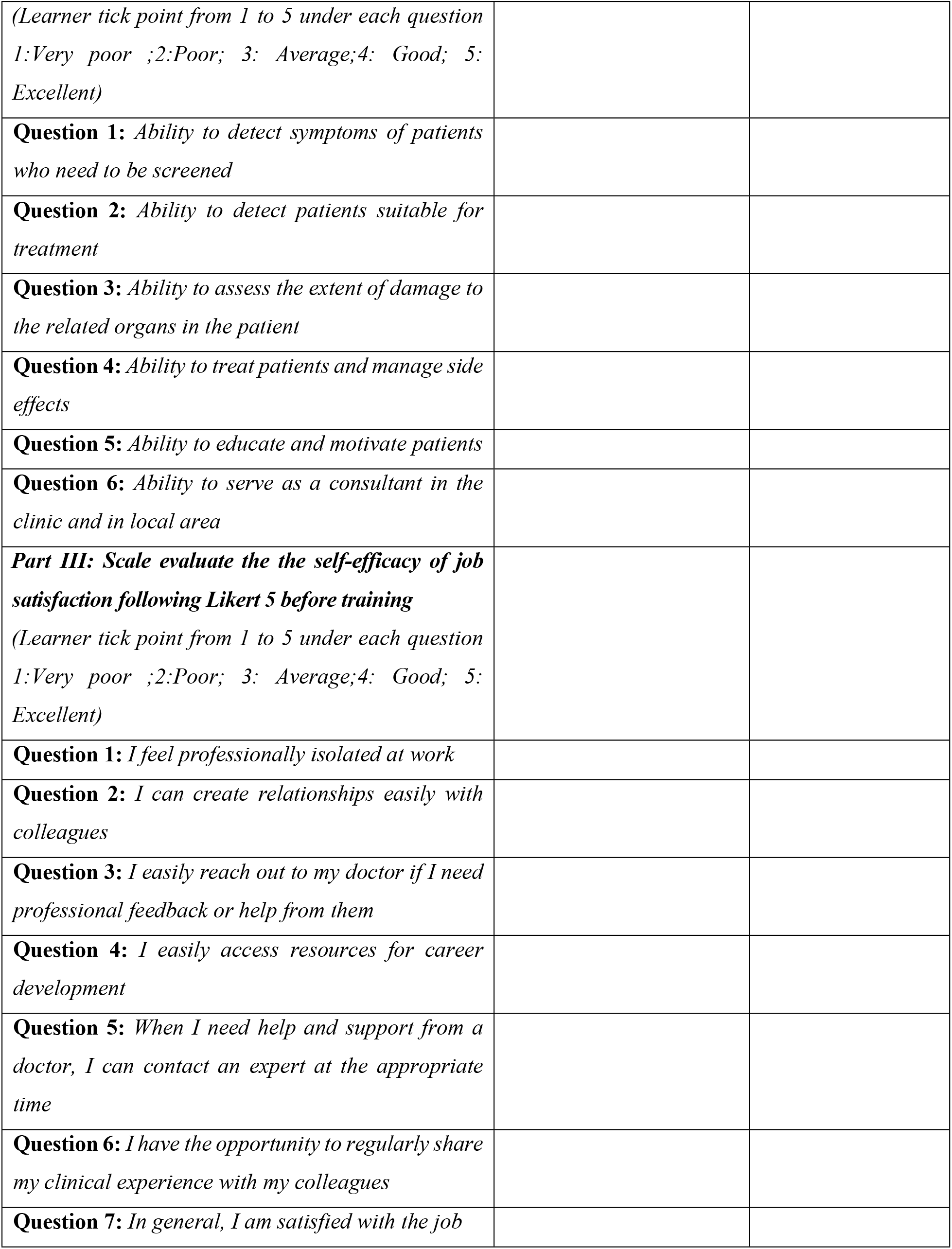
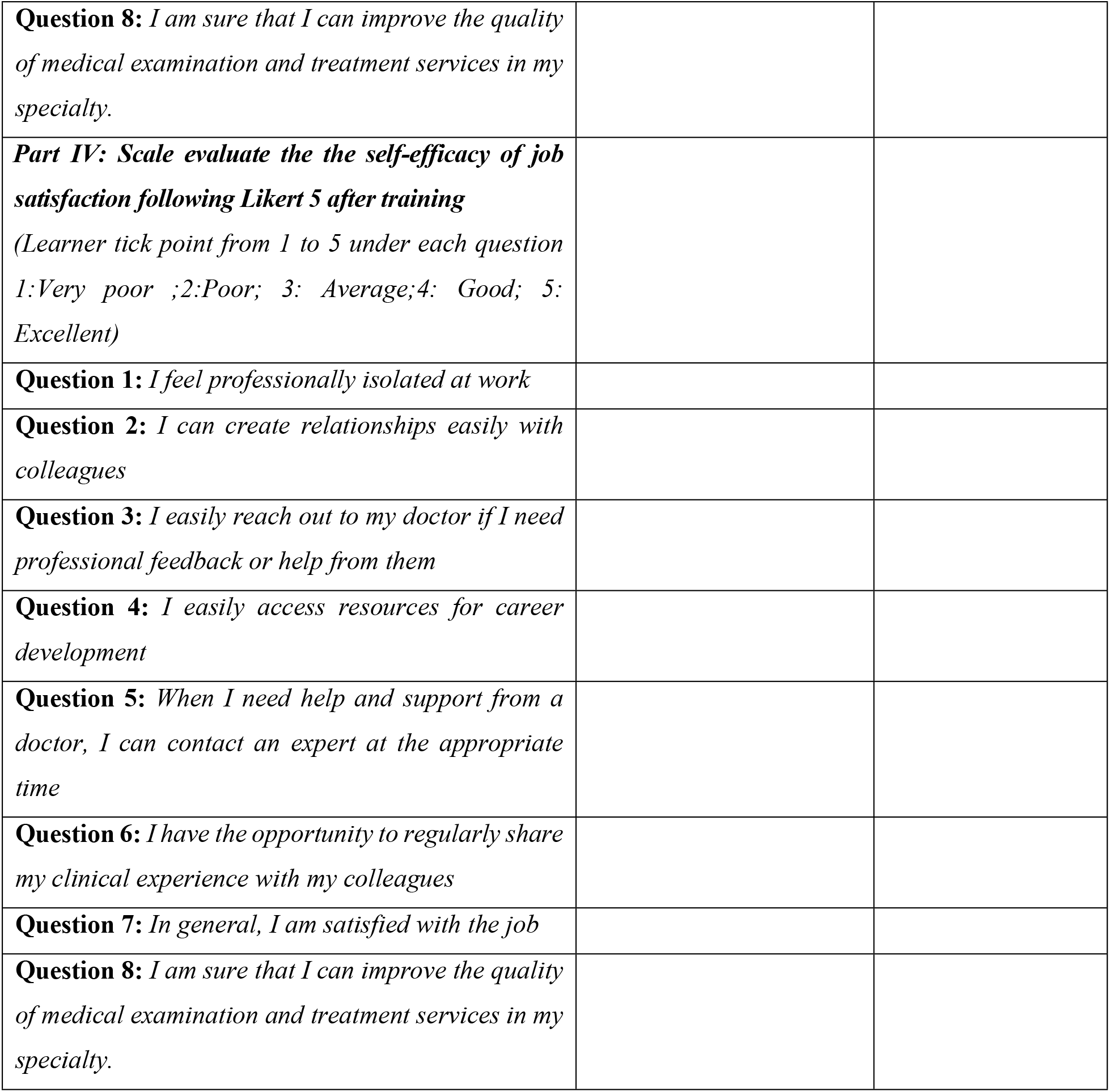

**"4 = very suitable"**
|  |  |  |
| --- | --- | --- |
| **Part I: Scale evaluate the the self-efficacy of knowledge, professional practice following Likert 5 before training**<br>*(Learner tick the point from 1 to 5 under each question 1:Very poor;2:Poor; 3: Average;4: Good; 5: Excellent)* | **PLEASE ASSESS THE APPROPRIATENESS OF THE SENTENCE S BY SCORING THEM ON EACH QUESTION**<br>*"1 = inappropriate"*<br>*"2= slightly appropriate"*<br>*"3= appropriate"*<br>*"4 = very appropriate"* | **Expert's comment in detail** |
| **Sentence 1:** *Ability to detect symptoms of patients who need to be screened* |  |  |
| **Sentence 2:** *Ability to detect patients suitable for treatment?* |  |  |
| **Sentence 3:** *Ability to assess the extent of damage to the related organs in the patient* |  |  |
| **Sentence 4:** *Ability to treat patients and manage side effects* |  |  |
| **Sentence 5:** *Ability to educate and motivate patients* |  |  |
| **Sentence 6:** *Ability to serve as a consultant in the clinic and in local area* |  |  |
| **Part II: Scale evaluate the the self-efficacy of knowledge, professional practice following Likert 5 after training** |  |  |

|  |
| --- |
| *(Learner tick point from 1 to 5 under each question*<br>*1:Very poor ;2:Poor; 3: Average;4: Good; 5:*<br>*Excellent)* |
| **Question 1:** *Ability to detect symptoms of patients*<br>*who need to be screened* |
| **Question 2:** *Ability to detect patients suitable for*<br>*treatment* |
| **Question 3:** *Ability to assess the extent of damage to*<br>*the related organs in the patient* |
| **Question 4:** *Ability to treat patients and manage side*<br>*effects* |
| **Question 5:** *Ability to educate and motivate patients* |
| **Question 6:** *Ability to serve as a consultant in the*<br>*clinic and in local area* |
| **Part III: Scale evaluate the the self-efficacy of job**<br>**satisfaction following Likert 5 before training**<br>*(Learner tick point from 1 to 5 under each question*<br>*1:Very poor ;2:Poor; 3: Average;4: Good; 5:*<br>*Excellent)* |
| **Question 1:** *I feel professionally isolated at work* |
| **Question 2:** *I can create relationships easily with*<br>*colleagues* |
| **Question 3:** *I easily reach out to my doctor if I need*<br>*professional feedback or help from them* |
| **Question 4:** *I easily access resources for career*<br>*development* |
| **Question 5:** *When I need help and support from a*<br>*doctor, I can contact an expert at the appropriate*<br>*time* |
| **Question 6:** *I have the opportunity to regularly share*<br>*my clinical experience with my colleagues* |
| **Question 7:** *In general, I am satisfied with the job* |

|  |
| --- |
| **Question 8:** *I am sure that I can improve the quality of medical examination and treatment services in my specialty.* |
| **Part IV: Scale evaluate the the self-efficacy of job satisfaction following Likert 5 after training**<br>(Learner tick point from 1 to 5 under each question<br>1:Very poor ;2:Poor; 3: Average;4: Good; 5: Excellent) |
| **Question 1:** *I feel professionally isolated at work* |
| **Question 2:** *I can create relationships easily with colleagues* |
| **Question 3:** *I easily reach out to my doctor if I need professional feedback or help from them* |
| **Question 4:** *I easily access resources for career development* |
| **Question 5:** *When I need help and support from a doctor, I can contact an expert at the appropriate time* |
| **Question 6:** *I have the opportunity to regularly share my clinical experience with my colleagues* |
| **Question 7:** *In general, I am satisfied with the job* |
| **Question 8:** *I am sure that I can improve the quality of medical examination and treatment services in my specialty.* |

### 3. Expert’s evaluation on scale

□ Experts assess whether the above items are appropriate to measure the "self-assessment of learners’ professional competence before and after participating in the Continuing Medical Education program”. This scale is applied for learners are health-providers working in the field of pediatrics.
□ Experts make any possible suggestions for adding or removing items or changing the wording of items on the scale.
□ Experts evaluate the instruction for the scale.
□ Experts evaluate the form of the scale.

